# Addressing Social Determinants While Social Distancing: The Development of an Evidence-Based Social Needs Screening for a Telehealth Setting

**DOI:** 10.1101/2021.10.23.21265399

**Authors:** Ruby E. Reed, María Valentina Suárez-Nieto, Jiwoo Lee, Neil K. Wary, Songnan Wang, Rachel Minseo Koo, Lars Osterberg, Wendy Caceres, Mina Charon, Amy Filsoof, Tamara Montacute, Baldeep Singh

**Affiliations:** Stanford University

**Keywords:** Social needs, Social determinants of health, Telehealth

## Abstract

Effectively addressing social determinants of health in clinical care can be challenging, and screening for such social needs is often overlooked. The COVID-19 pandemic has exacerbated health disparities and the impacts of social determinants of health, increasing the importance of both effective screening and intervention to address social needs. In response, the student-run free clinics at Stanford University sought to meet this need amongst our patient population by developing an evidence-based social needs screening (SNS) and referral protocol and integrating it into our novel telehealth model. The new protocol was implemented significantly more consistently compared to our previous checklist-based SNS, and more need was identified amongst our patient population than with the checklist-based, pre-pandemic screen. The new screening and referral protocol facilitated comprehensive patient care that addresses the social determinants of health in the clinical setting by improving our ability to identify patient social needs and refer such patients to community organizations. In describing the development, design, and implementation of this SNS, we hope to provide an example strategy for addressing social determinants of health within a student-run free clinic setting, and to encourage other student-run clinics and/or free clinics to similarly expand locally relevant social needs services.

## Introduction

Social determinants of health are non-medical factors that play a central role in shaping health outcomes. Such factors include socioeconomic status, education, social inclusion, and access to care, among many others.^1^ Underserved communities shoulder a disproportionate burden of unmet social needs.^2-4^ These disparities were exacerbated during the COVID-19 pandemic.^5-6^ The Stanford University Cardinal Free Clinics (CFCs) which consist of two student-run free clinics, Arbor Free Clinic (Arbor) in Menlo Park, CA and Pacific Free Clinic (PFC) in San Jose, CA, serve underserved and under-resourced patients: uninsured/under-insured, very low-income, many immigrants, and often non-English speaking. At the beginning of the pandemic, the CFCs closed for patient and volunteer safety.^7^ CFC staff immediately began developing a telehealth model.^8^ Given CFC patients were likely experiencing increased burdens of such needs during the pandemic,^9^ the CFC Community Outreach team sought to work with patients to address social needs alongside medical needs in this new care model.

Social needs screening (SNS) is a widely implemented strategy for care teams to identify unmet social needs for their patients. In a survey of 284 hospitals, 88% reported completing some type of social needs screening.^10^ Social needs screening generates useful referrals to community services in diverse clinical settings; however, there are also persistent challenges in the implementation of such screens, such as time, staffing limitations, and staff perceptions of screening.^11-13^

In 2018, the CFCs implemented a basic social needs checklist as part of the check-in process. This self-administered screen required patients to check boxes to indicate they had needs surrounding food insecurity, legal issues, health insurance, shelter, prescription assistance, utilities, and dental care. In 2019, the CFCs integrated a section into the electronic medical record (EMR) asking volunteers to indicate they had screened for domestic violence, substance use, and mental health concerns. Neither the checklist social needs screens nor the EMR items were developed from evidence-based practice, nor were they consistently implemented. Further, there was no formalized follow-up procedures for positive screens. Thus, these processes were not effective tools for addressing social determinants of health or understanding patient needs. Nevertheless, preliminary data collected from checklist screens and the EMR provided a snapshot of some of our patient populations’ needs.

Realizing the inadequacy of the current SNS protocol and the increasing urgency of social needs amidst the pandemic, the CFC Community Outreach team designed an evidence-based SNS and referral protocol based on existing literature, clinic data, knowledge of our patient population, and epidemiologic data from the counties we serve. After the SNS and referral protocols were developed, both CFCs adopted the new SNS and referral processes, with subjective success in identifying and referring for patient needs.

## Methods

### Evidence-Based Questionnaire Development

Initial development of the evidence-based SNS began in April 2020, and is visualized in **Figure 1**. Given the many social factors that affect health, and the limited time and resources available within each clinic visit, it was necessary to prioritize the needs most relevant to our communities. In this determination, CFC teams drew from three data sources: (1) data from our EMR and checklist-based social needs screens; (2) demographic and public health reports from the counties that most of our patients live in, so as to determine rates of common health and social issues such as unemployment, hunger, and other issues for which epidemiologic data was available; and (3) insights from research in similar populations or settings, such as primary care, obstetrics and gynecology, psychiatry, and emergency departments, examining typical needs addressed in care. Literature on social needs screening in these fields was reviewed to help determine which factors were deemed relevant to address within such settings. Identified social needs included: food insecurity, domestic violence, health insurance, housing, prescription assistance, substance use, utilities, and legal resources. A notable minority of our patients are visitors from other countries on temporary stays in the United States; we thus also elected to ask if the patient identifies as a non-immigrant “international visitor” (**Figure 1**).

**Figure 1.**
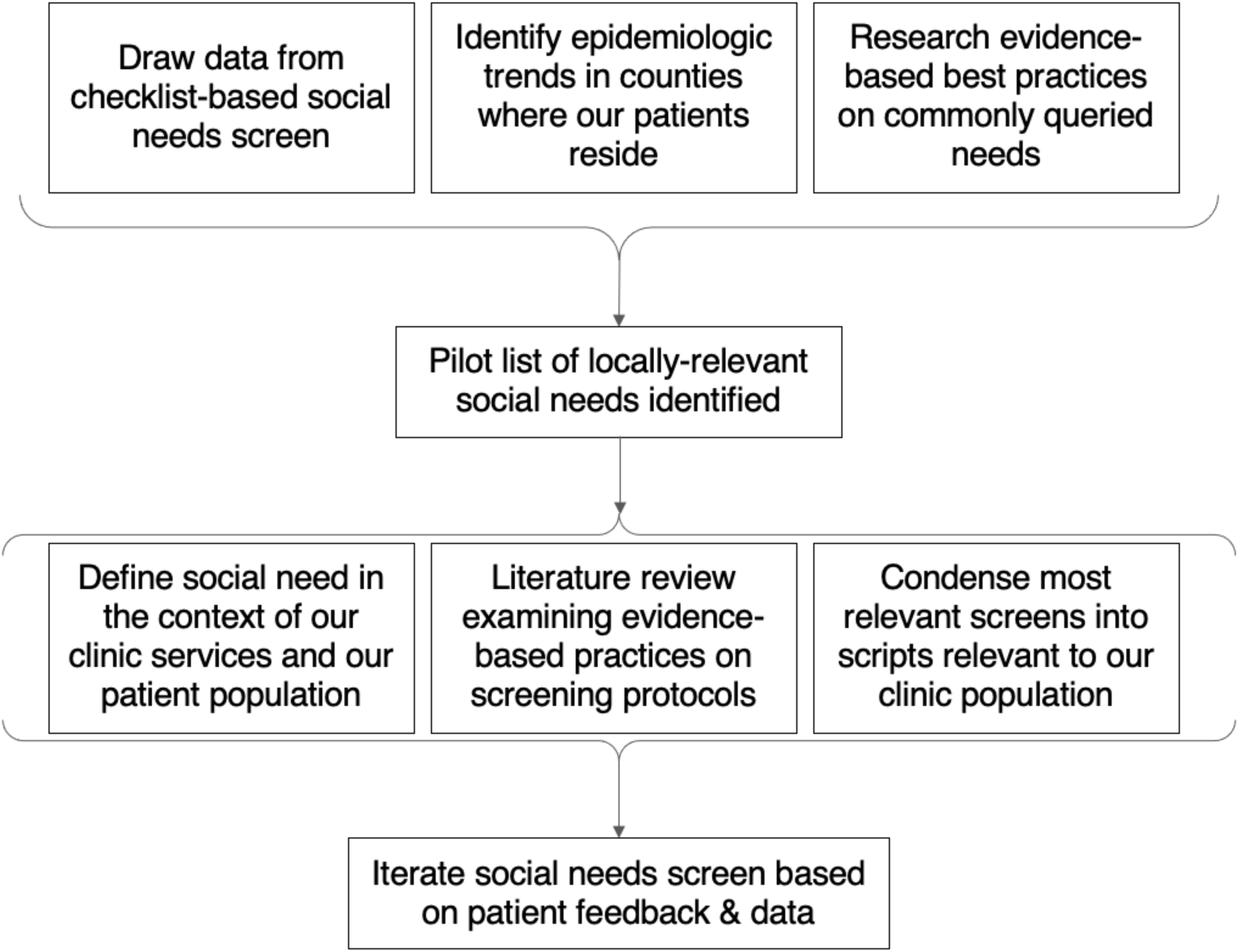
Development of evidence-based social need screen (SNS). To identify social needs, we utilized (1) our previous checklist-based EMR SNS, (2) epidemiologic and public health trends for our patient demographics, and (3) best practices on common queried needs in areas such as primary care and emergency medicine to identify social needs. After major social needs such as food insecurity, domestic violence, health insurance, housing, prescription assistance, substance use, utilities, and legal resources were identified, each social need was (1) defined in our free clinic context, (2) researched for evidence-based practices on how to ask question and screen, and (3) condensed and developed into scripts. Taken together, these steps formed the finalized evidence-based SNS.

Once social needs were identified, a screening protocol for each need was developed. Undergraduates taking part in a service learning course were each assigned a social determinant of their interest, and developed a screen for the related social need following a three-step process: (1) defining the social need in the context of the CFCs; (2) researching evidence-based screening practices for the need, with an emphasis on screening practices within clinical settings; and (3) condensing such practices and scripts to fit within a larger SNS that could be conducted in 10-15 minutes or less. Students were instructed to research evidence-based best practices using scholarly search engines (PubMed, Google Scholar), and recommendations from national and international government and medical organizations. Students presented their findings – highlighting screens/questions with the greatest evidence base, and suggesting modifications to fit the CFC setting. Clinic staff then condensed, synthesized, and incorporated a pilot SNS into the preliminary CFC telehealth flow (**Appendix 1**). All volunteers were trained on SNS implementation relevant to their roles using both instructional manuals and video scenarios of positive and negative screens created by the clinic management team.

### Referrals Protocol

Screening for social needs is of little utility without capacity to refer for services; thus, we concurrently expanded our social services search and referral capacity. We adopted a publicly available social service search engine to identify services and subsequently trained our referrals volunteers on its use. This search engine allows users to search for community organizations based on the type of social need, location, and certain identities or status factors, such as language, race, gender, age, or other factors that may affect eligibility. We also set up protocols and an information tracking system to ensure follow-up with patients was conducted in two weeks after initial visit.

### Implementation Within Telehealth Model

The CFC Telehealth Clinic launched on July 4, 2020, mirrors in-person offerings and consists of four student roles: front desk volunteers, who schedule appointments and register patients in the Electronic Medical Record (EMR); preclinical volunteers, who conduct the medical interview and report back to the attending physician; referrals volunteers; and patient health navigators (PHNs), who serve as a designated patient advocate and stay with the patient throughout their visit. This model and the additional innovations required for its implementation were described in further detail previously.^8^

Responsibilities for conducting the SNS are divided among three roles. Front desk volunteers conduct a pre-visit screening when scheduling patients. This includes SNS questions regarding linkages with a primary care physician, insurance status, need for assistance with prescription costs, and technology access. These needs are screened for prior to the patient visit to save time within the clinic day given their impact on referral planning processes.

Preclinical volunteers conduct the second part of the SNS as part of the initial medical interview. If not addressed elsewhere in the interview, the preclinical volunteer is instructed to administer the SNS surveys as part of the social history. The CFC EMR forms were restructured such that preclinical volunteers were prompted to conduct these screens using evidence-based scripts. The preclinical volunteers were tasked with assessing mental health, using the Patient Health Questionnaire-2 (PHQ-2)^14^ and Generalized Anxiety Disorder 2-item (GAD-2),^15^ expanding to the larger PHQ-9 and GAD-7 as indicated. A condensed version of the World Health Organization’s Alcohol, Smoking, and Substance Involvement Screening Test (ASSIST)^16^ was applied for substance use. If in initial review of substance use patients only endorsed use of alcohol or problematic alcohol use was suspected, volunteers were encouraged to screen with the Alcohol Use Disorders Identification Test (AUDIT).^17^ Further, preclinical volunteers provide education about domestic violence, as consistent with the Futures Without Violence Confidentiality, Universal Education and Empowerment, and Support (CUES) protocol.^18^ Direct query about experiences of domestic violence was not required, but preclinical volunteers were empowered to partner this education with a screen by asking directly about whether the patient is experiencing domestic violence. Volunteers were discouraged from indirectly asking about violence, such as by querying if the patient “feels safe at home”, due to the potential confusion this phrasing may cause, particularly during the pandemic, where patients may interpret this question to refer to safety from disease. PHNs recorded whether preclinical volunteers educated and/or screened patients, and which types of screens they used (i.e., direct vs. indirect). Throughout the patient’s encounter with the preclinical volunteer and attending physician, the PHN records all the patient’s social and medical referral needs in an internal form.

Once the medical interview is complete, the preclinical volunteer and attending physician leave the patient room to start planning the referrals for the patient with the labs and referrals coordination team. During this waiting time, the PHN conducts the third portion of the SNS, addressing food insecurity, utilities and housing security, legal needs, and employment assistance, among other needs. The SNS form prompts PHNs to reestablish safety and privacy and provides scripts for screening. As part of the SNS process, the PHN inputs patient needs from this final screening into the internal form. This form is immediately accessible to referral volunteers, who can integrate it into their plans for referrals for the patient. Referral volunteers then use the search engine to locate resources using the patients’ demographics and SNS data.

The protocol concludes with a warm hand-off script between the PHN and referral volunteer, who provides customized and comprehensive referral services for identified social and medical needs. Regardless of screening status, referrals volunteers conduct universal provision of locally-relevant information and resources surrounding staying safe from COVID-19, mental health, domestic violence, and basic needs such as food, employment, and housing. Referral information is sent to the patient with their after-visit summary via email or text, and two weeks after their visit, follow-up calls are made to confirm the patient has been able to access their referral services.

## Results

### Data Analysis

Data from the SNS from July 2020 through March 2021 and checklist-based SNS were pulled for the same period the previous year (July 2019-March 2020). Similar time periods were compared to reduce effects related to time of year. 505 unique patients were seen pre-pandemic during the months of July 2019 through March 2020. From July 2020 through March 2021, 181 unique patients were seen via telehealth and surveyed using the evidence-based SNS. For each patient, SNS questions regarding visitor status, utilities, mental health, legal needs, housing insecurity, domestic violence, and food were exported. Data were analyzed in R in counts of patient-questions, where 1 patient-question is equal to 1 question asked to 1 patient. Variables were compared using tests for differences in proportions, Welch Two Sample t-test, and 2-sample test for equality of proportions with continuity correction as specified. *p* < 0.05 was considered statistically significant.

### Demographics of Patients Seen In-Person and During Telehealth

We first hoped to understand if our telehealth population reflected our in-person population, as this may affect the applicability of our findings from the new data, and our ability to compare the checklist-based SNS to the current SNS. Notably, this is complicated by major gaps in the recording of demographic data within our clinics during the specified dates; this has been an area of ongoing quality improvement. **Table 1** describes the patient populations in our clinic seen from July 2019-March 2020 during in-person clinic as well as July 2020-March 2021 during telehealth. Data was pulled from the EMR in October 2021. For patients for which data was recorded, there is no significant difference between mean age (*p =* 0.5593), proportion of female patients (*p* = 0.1451), mean income (*p* = 0.5431), or ethnicity/race (*p* = 0.7085) in the telehealth versus the in-person clinic. A significantly higher proportion of telehealth patients used English as a first language (*p <* 0.001); however, significantly fewer of the in-person patients’ EMR includes reference to their spoken language (*p <* 0.001); it may be more likely that language is recorded when a translator is required.

**Table 1.** Demographic characteristics of patients from the Cardinal Free Clinics seen during July 2019 through March 2020 (in-person) compared to July 2020 through March 2021 (telehealth). Demographic characteristics of patients from the Cardinal Free Clinics seen during March 2019 through July 2020 (in-person) compared to March 2020 through July 2021 (telehealth). Reference date October 14, 2021. Mean age at reference date has not changed significantly (*P =* 0.5593) comparing in-person to telehealth using Welch Two Sample t-test. Biological sex has not changed significantly (*P* = 0.1451) using 2-sample test for equality of proportions with continuity correction. Language has changed significantly comparing in-person to telehealth (*P* = 9.092e-09) using 2-sample test for equality of proportions with continuity correction. Mean income did not change significantly (*P* = 0.5431) using Welch Two Sample t-test. Ethnicity/race did not change significantly (*P* = 0.7085) using 2-sample test for equality of proportions with continuity correction.

|  | July 2019 – March 2020<br>(n = 505)<br>n (%) | July 2020 – March 2021<br>(n = 142)<br>n (%) |
| --- | --- | --- |
| Mean age ± SD (years) | 50.01 ± 17.19 | 49.08 ± 16.78 |
| ≤ 30 years | 99 (19.6) | 26 (18.3) |
| 30 – 60 years | 212 (42.0) | 62 (43.7) |
| > 60 years | 194 (38.4) | 54 (38.0) |
| Biological sex |  |  |
| Female | 258 (51.1) | 83 (58.5) |
| Male | 216 (42.8) | 56 (39.4) |
| Not recorded | 31 (6.1) | 3 (2.1) |
| Language |  |  |
| English | 81 (16.0) | 55 (38.7) |
| Spanish | 41 (8.1) | 21 (14.8) |
| Mandarin | 56 (11.1) | 20 (14.1) |
| Cantonese | 2 (0.4) | 0 (0.0) |
| Vietnamese | 6 (1.2) | 1 (0.7) |
| Other | 31 (6.1) | 11 (7.7) |
| Not Recorded | 388 (57.0) | 34 (23.9) |
| Mean income ± SD (USD) | 29,093 ± 34,099.53 | 32,890 ± 6,228.57 |
| < \$25,000 | 24 (4.8) | 1 (0.7) |
| \$25,000 –\$50,000 | 12 (2.4) | 3 (2.1) |
| \$50,000 | 5 (1.0) | 0 (0.0) |
| Not recorded | 464 (91.9) | 138 (97.2) |
| Ethnicity/race |  |  |
| Asian | 79 (15.6) | 22 (15.5) |
| Hispanic/ Latinx | 43 (8.5) | 6 (4.2) |
| White | 12 (2.4) | 2 (1.4) |
| Other | 16 (3.2) | 5 (3.5) |
| Not Recorded | 355 (70.3) | 107 (75.4) |

### Success of New SNS Protocol

Using the new SNS, 2715 patient-questions were delivered, of which 1745 were answered (64.27%) (**Figure 2A**). This was significantly greater than the proportion of patients who were screened prior to telehealth using the prior SNS, of which 5952 patient-questions were delivered and 1618 were answered (27.18%) (*p* < 0.001) (**Figure 2B**).

**Figure 2.**
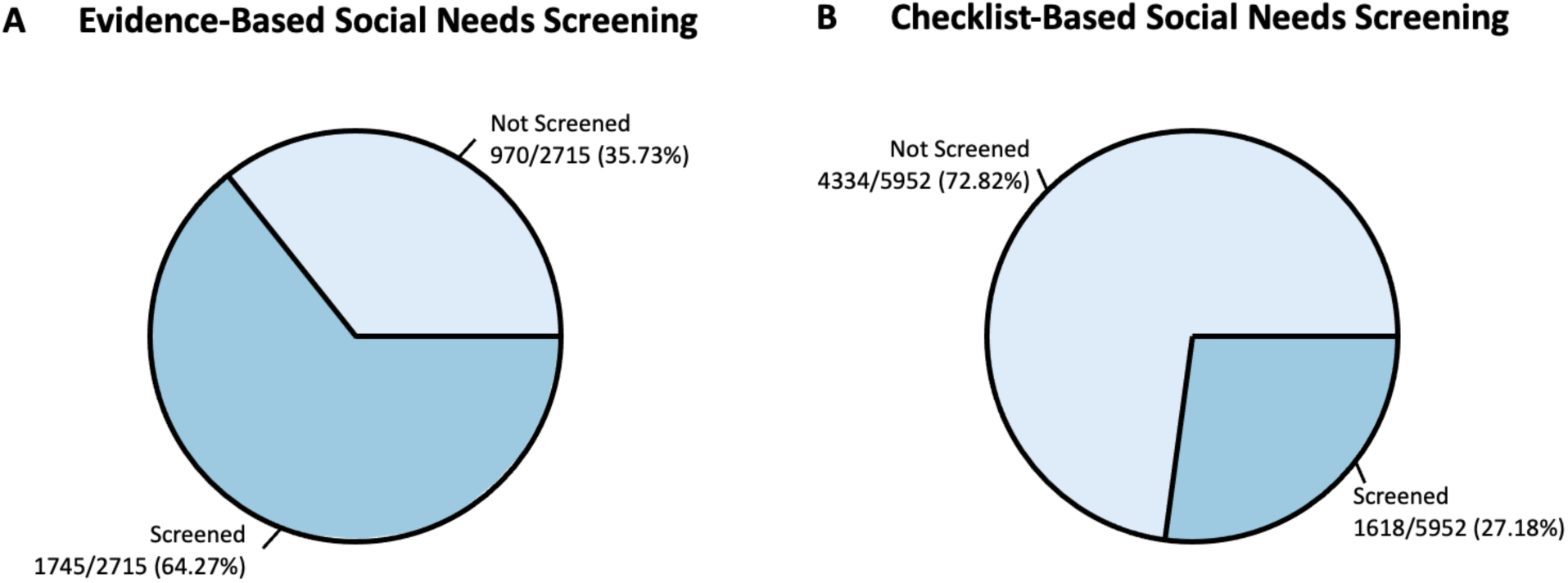
Percentage of screened patient-questions in the evidence-based SNS and checklist-based SNS. **(A)** Total number of possible patient-questions administered from the evidence-based social needs screening from July 2020 through March 2021. 1745 patient-questions were screened out of 2715 total possible opportunities (64.27%), while 970 patient-questions out of 2715 were omitted (35.73%). (**B**) Total number of possible patient-questions administered from the checklist-based social needs screening from July 2019 through March 2020. 1618 patient-questions were screened out of 5952 total possible opportunities (27.18%), while 4334 patient-questions out of 5952 were omitted (78.82%). The proportion of patient-questions administered in the evidence-based social needs screen was significantly (P < 0.001) greater than the proportion of patients-questions screened for social needs prior to telehealth using the checklist-based SNS.

The new SNS screens for visiting status, utilities, legal, housing, domestic violence, and food were implemented significantly more frequently (*p* < 0.001) (**Figure 3**). 52% of patients were not asked about their international visiting status – this may be because this status was apparent from the patient interview. The new protocol did not significantly improve screening rates for mental health issues (*p* = 0.310) (**Figure 3**). Although screening was improved, concerningly, 45% of patients were not screened or educated in any way about domestic violence under the new SNS.

**Figure 3.**
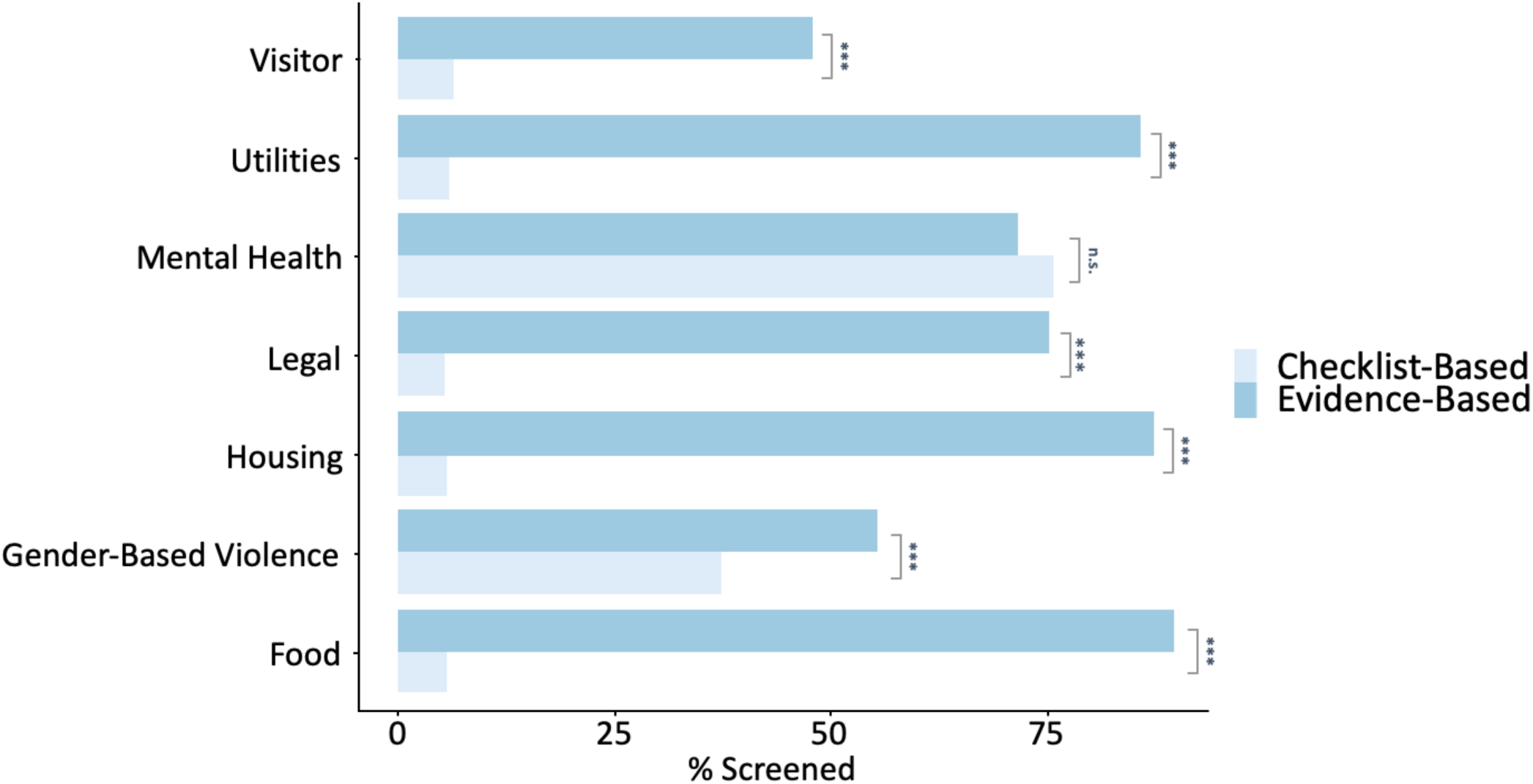
Comparison of % of patient-questions screened using the checklist-based SNS vs. the evidence-based SNS. Visitor status was screened 48% of the time using the evidence-based SNS while screened 6% of the time using the checklist-based SNS (*P* < 0.001). Utilities were screened 86% of the time using the evidence-based SNS while screened 6% of the time using the checklist-based SNS (*P* < 0.001). Mental health was screened 72% of the time using the evidence-based SNS while screened 76% of the time using the checklist-based SNS (*P* = 0.310). Legal needs were screened 75% of the time using the evidence-based SNS while screened 5% of the time using the checklist-based SNS (*P* < 0.001). Housing was screened 87% of the time using the evidence-based SNS while screened 6% of the time using the checklist-based SNS (*P* < 0.001). Gender-based violence was screened 55% of the time using the evidence-based SNS while screened 38% of the time using the checklist-based SNS (*P* < 0.001). Food insecurity was screened 90% of the time using the evidence-based SNS while screened 6% of the time using the checklist-based SNS (*P* < 0.001).

### Positive Screen of SNS for Different Types of Social Needs

Significantly more patients screened positive using the new SNS for requiring resources related to international visiting status, legal resources, housing, and food insecurity as compared to the old screen (*p* < 0.001 for all variables) (**Figure 4**). The proportion of positive screens was not significantly different for mental health (*p* = 0.764) and domestic violence (*p* = 0.736).

**Figure 4.**
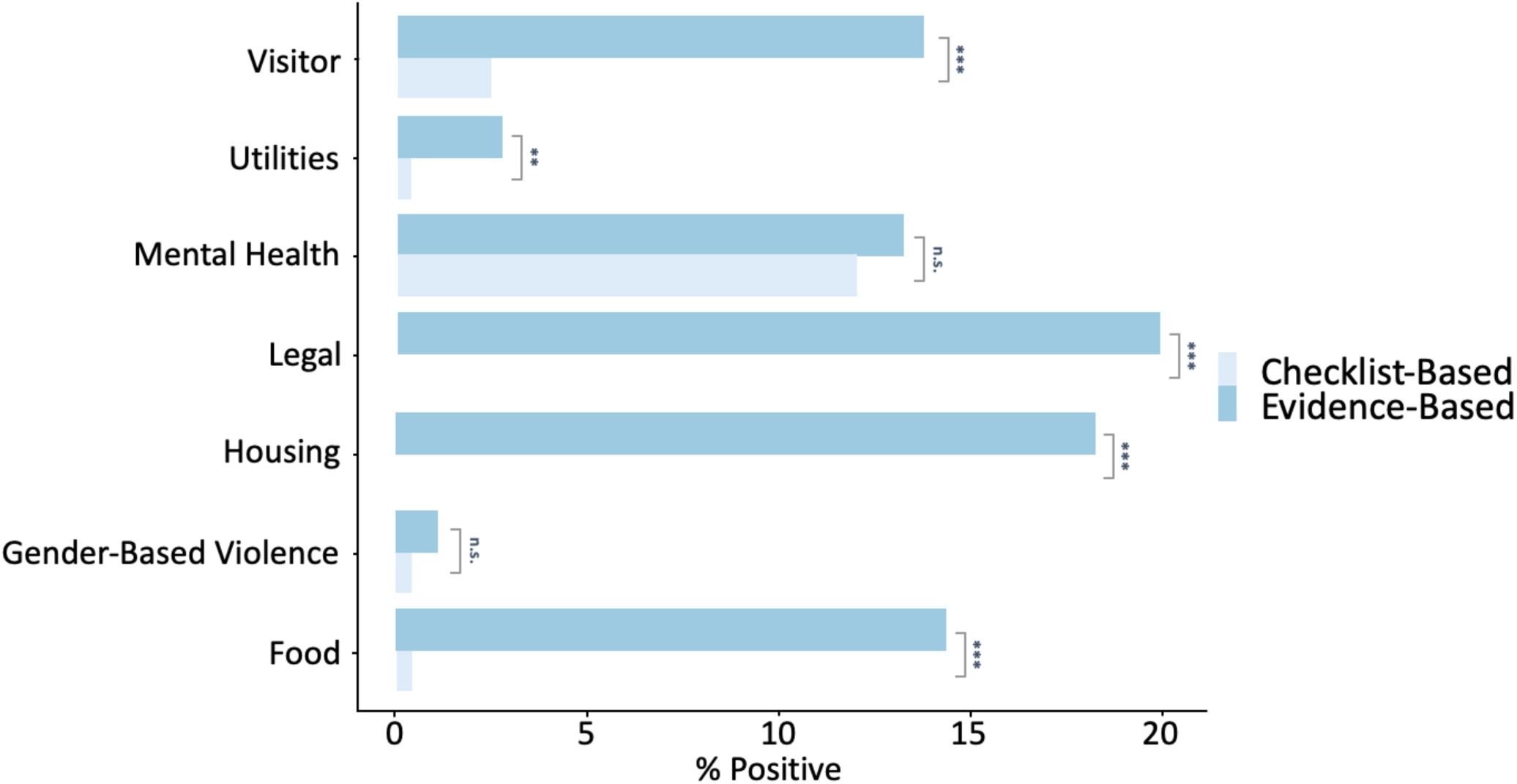
Comparison of % of positive patient-questions screened using the checklist-based SNS vs. the evidence-based SNS. Visitor status was screened positive 14% of the time using the evidence-based SNS while screened positive 3% of the time using the checklist-based SNS (*P* < 0.001). Utilities were screened positive 3% of the time using the evidence-based SNS while screened positive 0.4% of the time using the checklist-based SNS (*P* < 0.01). Mental health was screened positive 13% of the time using the evidence-based SNS while screened positive 12% of the time using the checklist-based SNS (*P* = 0.764). Legal needs were screened positive 20% of the time using the evidence-based SNS while screened positive 0.0% of the time using the checklist-based SNS (*P* < 0.001). Housing was screened positive 18% of the time using the evidence-based SNS while screened positive 0.0% of the time using the checklist-based SNS (*P* < 0.001). Gender-based violence was screened positive 1% of the time using the evidence-based SNS while screened positive 0.5% of the time using the checklist-based SNS (*P* = 0.736); however, it is notable that the goal of the evidence-based protocol was not disclosure. Food insecurity was screened positive 14% of the time using the evidence-based SNS while screened positive 0.4% of the time using the checklist-based SNS (*P* < 0.001).

## Discussion & Future Directions

The new SNS protocol has been successful in screening, identifying, and referring for social needs. However, no set of screening questions for social needs can be a static document, and we recognize our SNS as constantly iterative based on research, community needs, and community resources. Based on patient feedback, we have already incorporated more questions surrounding services for families with children and more detailed questions about legal and government services. However, we hope to utilize more rigorous methods to assess the comprehensiveness and acceptability of our social needs screening. We are thus currently conducting a study soliciting feedback from patients and community organizations on the SNS, and whether it adequately and accurately captures their needs. Results of this study will directly inform our SNS practices.

Better training of volunteers, delegation of clear roles and responsibilities, standardization of a protocol for screening, referral and follow-up, and the ability to track compliance through the completion of the SNS REDCap forms and EMR questions were likely all major contributors to the increase in screening rates under the new SNS protocol. The increased national conversations surrounding health equity and social determinants in light of the pandemic could also have encouraged screening. Patient engagement also likely played a role in screening success of the new SNS versus the old self-administered checklist SNS. Such checklists are time-saving for clinicians, but may leave patients with unanswered questions or unsure why they are being screened. Preliminary qualitative findings from our ongoing research supports the idea that our patients strongly support being asked explicitly about social needs by medical providers. As the CFCs transition to a hybrid in-person and telehealth model in October 2021, adherence to and patience satisfaction with the SNS should be monitored across both modalities, adapting for an in-person format.

Despite that the new SNS identified greater levels of need than the previous SNS, it is difficult to make meaningful comparisons about the needs of our patients pre- and post-pandemic due confounding variables, such as care delivery model (telehealth vs. in-person), potential changes in patient population, and the pandemic itself. Continued implementation of the SNS should reveal whether high need persists once pandemic restrictions are fully lifted. Regardless of its comparative screening success, we believe the new SNS does substantially better than its forebear at achieving its end-goal: helping the patient. Due to its apparent greater sensitivity to patient need, the new SNS helped our clinic to make substantially more referrals, connecting patients with resources during a time of heightened need.

Concerningly, our protocol has not significantly improved screening for mental health, substance use, or domestic violence. It is possible this reflects prior good practice; mental health and substance use screens were perhaps the most consistently implemented of all health screens prior to telehealth. However, reticence to screen may also be due to factors patient safety concerns, volunteer discomfort or lack of knowledge, time constraints, or stigma. Future investigations will examine the reasons that medical and physician assistant students skip certain screens. This research will guide training of volunteers to attempt to increase the screening for these sensitive but critical topics.

It is evident, now more than ever, that health care organizations must assess and address social determinants of health in a routine, evidence-based manner. There are major barriers to screening and referral in clinical settings, particularly in those serving underserved and under-resourced patients. However, such limitations should not prevent important questions from being asked. We hope our efforts encourage further innovation to incorporate comprehensive, locally relevant SNS protocols into workflow in other clinical settings. As we examine and iteratively improve our SNS, we implore other clinics serving the underserved to join us in working towards our goals: greater patient satisfaction and empowerment, more successful referrals, and a greater ability of our clinic to treat the whole person and the whole community.

## Data Availability

All data produced in the present study are available upon reasonable request to the authors.

## Acknowledgments

We would like to thank the contributions of the many volunteer physicians and students who help organize our clinics.

## APPENDIX 1: Social Needs – Screening Questionnaire

### Part 1. Front Desk Pre-Visit Screening

#### Insurance Status

A. We do not require insurance for you to be seen at the clinic, and we are a free service. However, we would like to know your insurance status so we can best assist you in achieving your health goals. Do you have insurance?
  - **If the patient answers “Yes”**, proceed to “Primary Care Provider” section
  - **If the patient answers “No”**, proceed to asking B.
B. Insurance can help with affording medical care. We have resources on how to get low or reduced cost insurance. Are you interested in learning more about how to get insurance?
  - **If the patient answers “Yes”**, proceed to referring them to B2C for insurance counseling
  - **If the patient answers “No”**, proceed to “Primary Care Provider” section

#### Primary Care Provider

A. Do you currently have a primary care provider?
  - **If the patient answers “Yes”**, proceed to asking, “When did you last visit your primary care doctor?”
  - **If the patient answers “No”**, proceed to asking B.
B. We have resources on how to connect with primary care providers. Are you interested in learning more about how to get insurance?
  - **If the patient answers “Yes”**, proceed to referring them to B2C for referral to a primary care provider.
  - **If the patient answers “No”**, proceed to “Prescription Assistance” section

#### Prescription Assistance

A. Do you currently take any medications?
  - **If the patient answers “Yes”**, proceed to asking B.
  - **If the patient answers “No”**, proceed to asking C.
B. Does the cost of your medication concern you?
  - **If the patient answers “Yes”**, proceed to asking C.
  - **If the patient answers “No”**, proceed to concluding the screening questionnaire.
C. We have some resources that may help you to reduce the cost of your medications. Would you be interested in learning more about these resources during your visit?
  - **If the patient answers “Yes”**, proceed to referring them to B2C for prescription assistance counseling.
  - **If the patient answers “No”**, proceed to concluding the screening questionnaire.

##### Results

If the patient screens positive for any of the above, refer them to the appropriate insurance, primary care provider, or prescription assistance resources via B2C or referral program.

### Part 2. Pre-clinical Volunteer Screening

#### Safety and Privacy

State: “Your medical information is confidential, and that doesn’t change just because you’re not in a physical health center setting. I will not share anything we talk about here outside of the care team.”

A. Are you somewhere where you feel safe to speak privately?
  - **If the patient answers “Yes”**, proceed to offer both mental health and substance use training, as well as universal education about violence and health, as part of social needs screening.
  - **If the patient answers “Yes” and is with family members and/or friends as part of the visit**, proceed with social needs screening, including mental health and substance use screening, but DO NOT conduct universal Gender Based Violence (GBV) education. Resources for GBV will still be provided, if patient consents, via B2C or referrals program.
  - **If the patient answers “No”, but is alone**, work with the patient to support their safety and privacy during telehealth visit by considering 1-3. Allow the patient time to adjust accordingly and proceed to 4.
    1. Ask: “Are you able to move to a place where you feel more comfortable to talk freely?”
    2. Ask: “Would you prefer to find another time to talk?”
    3. Work to understand the privacy situation. As relevant, suggest alternatives:
      ▪ Alternative location (i.e. move to bathroom, car, etc.)
      ▪ Alternative device (i.e. move to phone vs. laptop or iPad)
      ▪ Headphones for patient or for nearby children
    4. Ask again: “Are you somewhere where you feel safe to speak privately?”
      · **If the patient answers “Yes”**, proceed with social needs screening, including GBV universal education.
      · **If the patient answers “No”**, proceed with social needs screening, including mental health and substance use screening, but DO NOT conduct universal GBV education. Resources for GBV will still be provided, if patient consents, via B2C or referrals program.

#### Women’s Health Maintenance

A. Ask if the patient identifies as a woman.
  - **If the patient answers “Yes”**, proceed to B and C.
  - **If the patient answers “No”**, proceed to “Mental Health” Section

2. When was your last mammogram?
  - If the patient is over 50 and states that their last one was more than 2 years ago, proceed to referring them to B2C or referrals program for mammogram referral.
  - Otherwise, proceed to C.

3. When was your last pap smear?
  - If the patient is over 21 and states that their last one was more than 3 years ago, proceed to referring them to B2C or referrals program for pap smear referral.
  - Otherwise, proceed to “Mental Health” Section

#### Mental Health

A. Current events have caused a lot of changes for many people, and many people are feeling stressed, low or anxious. Over the last two weeks, how often have you been bothered by…
  1. Having little interest or pleasure in doing things? **Select one of the following based on patient’s response:**
    - Not at all
    - Several days
    - More than half the days
    - Nearly everyday
  2. Feeling down, depressed, or hopeless? **Select one of the following based on patient’s response:**
    - Not at all
    - Several days
    - More than half the days
    - Nearly everyday
  3. Feeling nervous, anxious, or on edge? **Select one of the following based on response:**
    - Not at all
    - Several days
    - More than half the days
    - Nearly everyday
  4. Not being able to stop or control worrying? **Select one of the following based on response:**
    - Not at all
    - Several days
    - More than half the days
    - Nearly everyday

#### Substance Use

A. Which one of the following substances have you ever used in your lifetime?
  - Alcohol
  - Cigarettes
  - E-cigarettes, vapes, or Juul
  - Marijuana
  - Opioids or prescription painkillers (ex., fentanyl, hydrocodone, oxycodone, buprenorphine, methadone)
  - Methamphetamine or other amphetamines
  - Heroin or other injectable drug
  - Cocaine or crack cocaine
  - Ecstasy or MDMA (“Molly”)
  - Inhalants (nitrous, glue, “poppers”)
  - Hallucinogens such as LSD, psilocybin (mushrooms), ayahuasca
  - Other: (If the patient indicates that the drug used is not listed, please note name of drug next to **‘Other’**.) **If the patient screens positive** for any of the above substances, proceed to asking B-H For each substance mentioned. **If the patient screens negative** for all substances, and a safe and private location was confirmed from the “Safety and Privacy” section, proceed to “Gender-based Violence” section. Otherwise, conclude screening.
B. In the past 3 months, how often have you used each of the substances you mentioned [first drug, second drug, etc.]? **Select one of the following based on response:**
  - Never
  - Once or Twice
  - Monthly
  - Weekly
  - Daily or Almost Daily
  **If the patient answers “Never”** skip to F. Otherwise, continue to C-H.
C. In the past 3 months, how often have you had a strong desire or urge to use? **Select one of the following based on response:**
  - Never
  - Once or Twice
  - Monthly
  - Weekly
  - Daily or Almost Daily
D. In the past 3 months, how often has your use of [first drug, second drug, etc.] led to health, social, legal, or financial problems? **Select one of the following based on response:**
  - Never
  - Once or Twice
  - Monthly
  - Weekly
  - Daily or Almost Daily
E. In the past 3 months, how often have you failed to do what was normally expected of you because of your use of [first drug, second drug, etc.]? **Select one of the following based on response:**
  - Never
  - Once or Twice
  - Monthly
  - Weekly
  - Daily or Almost Daily
F. Has a friend or relative or anyone else ever expressed concern about your use of [first drug, second drug, etc.]? **Select one of the following based on response:**
  - No
  - Yes, but not in the past three months
  - Yes, in the past three months
G. Have you ever tried and failed to control, cut down, or stop using [first drug, second drug, etc.]? **Select one of the following based on response:**
  - No
  - Yes, but not in the past three months
  - Yes, in the past three months
H. Have you ever used any drug by injection? (nonmedical use only) **Select one of the following based on response:**
  - No
  - Yes, but not in the past three months
  - Yes, in the past three months

#### Gender-based Violence

A. Before we get started, I want to say that I know COVID-19 has made things harder for everyone. Because people are stressed, we are sharing ideas about helping yourself and people you care about. For example, we may experience more stress now in our relationships including increased fighting or harm, and that can affect our health. There is free, confidential help available if you know someone who is being hurt in their relationship. Would it be okay if I sent some resources for you to share?
  - **If the patient answers “Yes”**, refer to B2C for designated all-patient GBV resources, even if there is no disclosure of violence, exploitation, or relationship troubles and proceed to B.
  - **If the patient answers “No”**, proceed to B.
B. How are things going right now for you?
  - **If no disclosure**, affirm patient experiences and feelings. Move to next portion of clinical interview Even if patient is not currently experiencing violence or exploitation, they may still be feeling triggered or in crisis because of COVID-19 situation.
    · Validate these feelings
    · Offer strategies for reducing stress *(can offer Bridge 2 Care referral for wellness resources)*
    · Answer questions about COVID-19, and staying healthy in the pandemic
    · Connect to support services for mental health, food access, other needs, etc.
    **If disclosure** (**positive screen)**, complete the following based on situation:
  1. If patient discloses a current injury related to violence: **In this situation, mandatory reporting is required**.
    · Safely and considerately end interview ASAP. Explain you will be back with the physician.
    · Tell MD, manager and B2C of +GBV screen.
    · Bring MD into room with patient.
    1. If patient discloses current violence or exploitation experiences without current injury, or indicates that things with their relationship are difficult or stressful, offer validating messages and a warm referral to an GBV support agency, crisis text line, and health promotion information
    2. Thank you for sharing this with me. I am so sorry this is happening. It is not right, and it is not your fault. What you are telling me makes me worry about your safety and health. A lot of our patients experience things like this, and there are people who can help. I can connect you today with some resources that may help. We can work together with our referral coordinator to figure out a way to deliver these resources to you safely. How does that sound?
      · **If the patient agrees or says “Yes”**, proceed to C.
      · **If the patient does not agree or says “No”**, conclude interview
C. Ask about effects on health maintenance and COVID-19 safety, as this may affect care plan: “These issues can affect our health in many ways. I’d like to hear if your partner or someone else is interfering in any way with your plans to stay healthy during this time. This may include disrupting your ability to take your medicines, taking away hand sanitizer, preventing you from seeking help, or keeping you from connecting with friends and family.”
  - In your summary statement to patient, mention that you will be bringing this up with the physician.
  - Ensure that you mention the +GBV screen to the physician, as it likely will affect care planning.
  - Accompany patient to B2C to ensure warm GBV organization referral

##### Results

If patient screens positive for either mental health, substance use screening, or GBV, refer to necessary clinical resources via B2C or referral program. Refer to Bridge to Care for GBV services, with positive indication for GBV (safe resource delivery).

### Part 3. Patient Health Navigator Social Needs Screening

**PHN:** “This is a stressful time for many people, and it is important we all protect ourselves from COVID-

At the end of this visit, we have some resources that we will provide you with about how to maintain your health.”

**PHN:** “However, beyond just preventing illness, in this time, many people are having a hard time paying for things they need, or are facing a lot of extra stress. We are asking all our patients a few questions about these types of needs to get a better understanding of how you’re doing and how we can be of better help to you. Would it be all right if I ask you a few questions about these types of issues?”

#### Food Insecurity

A. Many people during this time are experiencing challenges in finding and affording healthy food. Within the past month, have you or your family worried that your food would run out before you got money to buy more? **Select one of the following based on response:**
  - Yes
  - Sometimes
  - No
  - Don’t know or refuse to answer
B. Within the past month, have you or your family noticed that the food you bought just didn’t last and you didn’t have money to get more? **Select one of the following based on response:**
  - Yes
  - Sometimes
  - No
  - Don’t know or refuse to answer

**Legal (Working conditions/Employment Issues/Immigration)**

*Note: Skip this section if the patient is a visitor to the U*.*S*.

A. The COVID-19 pandemic has affected employment for many people. Are you currently employed?
  - **If the patient answers “Yes”**, ask: Are you having any issues with your job or employment status such as not receiving proper pay or unsafe working conditions?
  - **If the patient answers “No”**, ask: Are you interested in seeing your eligibility for unemployment benefits/compensation?

2. Are you aware of benefits you may be eligible for, such as Medicare, Medicaid or Disability benefits?
  - **If the patient answers “Yes”**, proceed to C
  - **If the patient answers “No”**, ask: Are you interested in seeing your eligibility for unemployment benefits/compensation?
3. Have you applied for these programs and received those benefits?
  - **If the patient answers “Yes” and received benefits**, proceed to E
  - **If the patient answers “Yes” and did not receive all benefits**, ask: Do you know why you were unable to receive those benefits?”
  - **If the patient answers “No”**, proceed to D

D. Would you be interested in learning more about how to apply for the benefits?
  - **Record patient’s response** and proceed to E

5. Legal services can be very helpful for people and families who are working to establish a safe immigration status. Some of these services may be free or low-cost. Would you like to receive information about legal resources related to immigration?
  - **If the patient answers “Yes”**, refer to B2C and proceed to “Utilities/Housing” section
  - **If the patient answers “No”**, proceed to “Utilities/Housing” section

#### Utilities/Housing

A. Are you and your family concerned that in the next two months, you may not have stable housing that you own, rent, or stay in as a part of a household?”
  - Yes
  - No
B. “In the past 12 months has the electric, gas, oil, or water company threatened to shut off services in your home?”
  - Yes
  - No
  - Already shut off

##### Results

Patients screen positive for food insecurity if the response is “yes” or “sometimes” for either or both statements. Refer to Bridge to Care for resources in helping afford, government benefits, nutrition education, emergency food, food delivery, food pantry, meals. If a patient is unemployed, in need of benefits, or has applied for benefits they did not receive, it is considered a positive screen. Refer to appropriate legal and employment resources via B2C. If a patient screens positive by answering “yes” and/or “Already shut off” for utilities and/or housing, refer to housing resource and/or resource to help pay for utilities via Bridge to Care.

